# Natural language processing and expert follow-up establishes tachycardia association with *CDKL5* deficiency disorder

**DOI:** 10.1101/2023.06.24.23291719

**Authors:** Alina Ivaniuk, Christian M Boßelmann, Xiaoming Zhang, Mark St John, Sara C Taylor, Gokul Krishnaswamy, Alex Milinovich, Peter F Aziz, Elia Pestana-Knight, Dennis Lal

## Abstract

**Purpose:** CDKL5 deficiency disorder (CDD) is a developmental and epileptic encephalopathy with multisystemic comorbidities. Cardiovascular involvement in CDD was shown in animal models but is yet poorly described in CDD cohorts.

**Methods:** We identified 38 individuals with genetically confirmed CDD through the Cleveland Clinic CDD specialty clinic and matched 190 individuals with non-genetic epilepsy to them as a comparison group. Natural language processing was applied to yield Human Phenotype Ontology (HPO) terms from medical records. We conducted HPO association testing and manual chart review to explore cardiovascular comorbidities associated with CDD.

**Results:** We extracted 243,541 HPO terms from 30,512 medical encounters. Phenome-wide analysis confirmed well-established CDD phenotypes and identified association of tachycardia with CDD (OR 4.2, 95%CI 1.75-9.93, p_adj_<0.001). We found a 99.6-fold enrichment of supraventricular tachycardia (SVT) in CDD encounter notes (p_adj_ < 0.001), which led to identification of two cases of fetal/neonatal onset SVT previously undescribed in CDD. Tachycardia in CDD individuals was associated with the presence of other autonomic symptoms (OR 5.63, 95%CI 1.08-40.3, p=0.038).

**Conclusions:** CDD is associated with tachycardia, potentially including early-onset supraventricular tachycardia. Alongside prospective validation studies, semiautomated genotype-phenotype analysis with matched controls is a scalable, rapid, and efficient approach for validating known and identifying novel phenotype associations.

## Introduction

CDKL5 deficiency disorder (CDD; MIM:300572, MONDO:0010396) is a rare genetic disease caused by pathogenic variants in the *CDKL5* gene coding for cyclin-dependent kinase-like 5 protein. CDD primarily affects the central nervous system resulting in early-onset pharmacoresistant epilepsy, developmental delay, sleep disturbances, hypotonia, and visual cortical impairment.^1^ Other systems are also involved in CDD resulting in a range of comorbidities, including gastrointestinal symptoms, sleep disorders, scoliosis, and respiratory issues.^2 3^

Cardiovascular comorbidities in CDD represent an area of ongoing research.^14^ Cardiovascular phenotypes, in particularly long QT syndrome, is well-described in Rett syndrome, a clinically related disorder.^5^ Recently, QT interval prolongation and tachycardia were observed in a mouse model of CDD^6^, but it is yet unclear if these cardiac findings translate to humans. Current evidence regarding cardiovascular involvement in CDD is limited two studies with inconsistent findings. A caregiver survey that found 11 out of 29 individuals with CDD who underwent electrocardiogram (ECG) to have arrhythmia.^3^ Another recent clinical cohort study reported a low prevalence of long QT syndrome in their CDD cohort^7^. As a result, the prevalence and clinical relevance of the full spectrum of cardiac symptoms in a real-world cohort of individuals with CDD remains poorly understood, which complicates development of care recommendations for individuals with CDD.^8^

Studying comorbidities in rare diseases is challenging due to a limited number of cases and clinical information. New digital approaches, such as natural language processing and electronic health records (EHR) research, can facilitate characterization of disease associations.^9,10^ To address the existing knowledge gap about cardiovascular involvement in CDD, we performed an EHR-based deep computational phenotyping of 38 individuals with CDD with a comparative analysis of comorbidities against a cohort of 190 matched individuals with epilepsy receiving care within a single healthcare network.

## Methods

### 1. Study cohort construction CDD cohort acquisition

In this retrospective study, we used clinical data of patients with CDD receiving care at the Cleveland Clinic Foundation (CCF) *CDKL5* Center of Excellence endorsed by the International Foundation for CDKL5 Research. The initial cohort included patients from an in-house *CDKL5* patient database (n=42). We additionally queried the electronical health record (EHR) database using the ICD-10 code for Cyclin-Dependent Kinase-Like 5 Deficiency Disorder (G40.42, n=32) and screened the resulting 50 unique medical records for variants in *CDKL5* gene. We included 38 patients with pathogenic variants in *CDKL5* gene (Supplementary Figure 1).

#### Comparison cohort acquisition

Since childhood-onset epilepsy is an early and universal feature of CDD^11^, we chose individuals with childhood-onset epilepsy as a comparison cohort. Based on the most comprehensive to-date systematic review on the accuracy of approaches to identify epilepsy using administrative healthcare data^12^, we used the following criteria to identify individuals with epilepsy in the EHR database: a) Having an International Classification of Diseases, Tenth Revision, Clinical Modification (ICD-10-CM) code G40 (“Epilepsy and recurrent seizures”) or ICD-9-CM code 345.*; b) Having a Current Procedural Terminology (CPT) code for any type of electroencephalography (EEG); c) Age 0-5 years at time of diagnosis (defined as an age of the first billing code for epilepsy). We used a stricter definition to reduce diagnostic heterogeneity and increase enrichment of the resulting cohort for epilepsy.

To ensure encounter data homogeneity across groups, we matched individuals with CDD to control individuals using the following variables: age at encounter, sex, ethnicity, and number of encounters. We used a 1:5 variable ratio, parallel, balanced nearest neighbor propensity score matching estimated by a generalized linear model.

#### EMR-based screening of phenotypic profiles of CDD against comparison cohort

##### Clinical variable extraction

The clinical variables from patient encounters for the study were extracted from the CCF Research Data Warehouse (RDW). The RDW represents an in-house relational database that utilizes concepts from Unified Medical Language System (UMLS, release 2022AA), a thesaurus that integrates different biomedical vocabularies, allows for standardization of health medical records, and facilitates use of biomedical information in research. The RDW development has been previously described^13^ and is summarized in Supplementary Figure 2.

We queried the annotated encounter data for the identified CDD and epilepsy cohort and mapped the UMLS concepts to Human Phenotype Ontology (HPO, v.2023-01-27) terms. HPO is a hierarchical vocabulary of phenotypic features specifically designed to study rare genetic disorders.^14^ We then performed HPO term propagation to higher-level clinical concepts to enable broader phenotypic characterizations and comparisons (Supplementary Figure 3).^15^ Age at encounter, age at last follow-up, and age at diagnosis were calculated in relationship to the date of birth based on encounter dates. While focusing on cardiovascular features, we also analyzed other phenotypes to explore whether established CDD phenotype associations can be identified in a computational analysis of real-world data, informing accuracy of this hypothesis-free approach.

##### Computational analysis

We used ratios, medians and interquartile ranges (IQR) for descriptive statistics. To compare phenotypic features between CDD and non-genetic epilepsy patients, we applied Fisher’s exact test with Bonferroni correction for multiple testing at a significance level of α=0.05. We reported the effect size of relative enrichment as odds ratio with 95% confidence intervals (CI). Statistical analyses and cohort matching were performed using the R programming language (v. 4.2.0).

#### 3. Case review

EHRs of the CDD patients were reviewed to verify the identified phenotype associations and to extract additional information to concisely summarize the clinical findings. Available electrocardiograms (ECGs) were reviewed by a board-certified pediatric cardiologist.

## Results

We identified 38 individuals with CDD for the study. The unmatched epilepsy cohort included 1671 individuals. We extracted demographic and encounter data from 182,919 medical encounters to identify a subset of 190 individuals with epilepsy matching the CDD cohort (Supplementary Table 1). The median length of follow-up for the CDD and matched cohorts was 8.4 (IQR 2.88-18.35) years with a cumulative follow-up of 2,525 person-years. The median age at the first and last encounter were 0.2 (IQR 0.0-1.1) and 11.6 (IQR 6.5-16.3) years for CDD and 0.2 (IQR 0.0-1.3) and 9.1 (IQR 4.9-19.1) for the comparison cohort, respectively. A total of 227,370 UMLS concepts across 12,699 encounters mapped to 298,913 propagated HPO terms were available for analysis.

We first explored the individual frequencies of unspecific HPO terms describing body systems abnormalities. We found that abnormality of digestive system physiology (HP:0025032) and abnormal cardiovascular system physiology (HP:0011025) are encountered statistically significantly more frequently in CDD than in comparison cohort (p_adj_=0.03 for both; Figure 1A). We then analyzed relative enrichment of individual occurrence of children HPO terms of these two terms, along with abnormal nervous system physiology (HP:0012638), which is the core phenotype in CDD, and abnormality of eye (HP:0000478), which we suspected to show the signal in CDD because of cerebral visual impairment (HP:0100704). We found a significant enrichment for known CDD comorbidities^2^ as well as for tachycardia (HP:0001649; OR 4.2, 95%CI 1.75-9.93, p_adj_<0.001; Figure 1B). Encounter-based analysis of HPO terms showed, in addition to enrichment for terms denoting a known CDD phenotype, enrichment of tachycardia (HP:0001649; OR 3.22, 95%CI 2.51-4.12, p_adj_<0.001), as well as a significant enrichment for supraventricular tachycardia (OR 99.6, 95%CI 43.79-278.59, p_adj_<0.001), a finding that has not been yet specified for CDD (Figure 1C).

**Figure 1.**
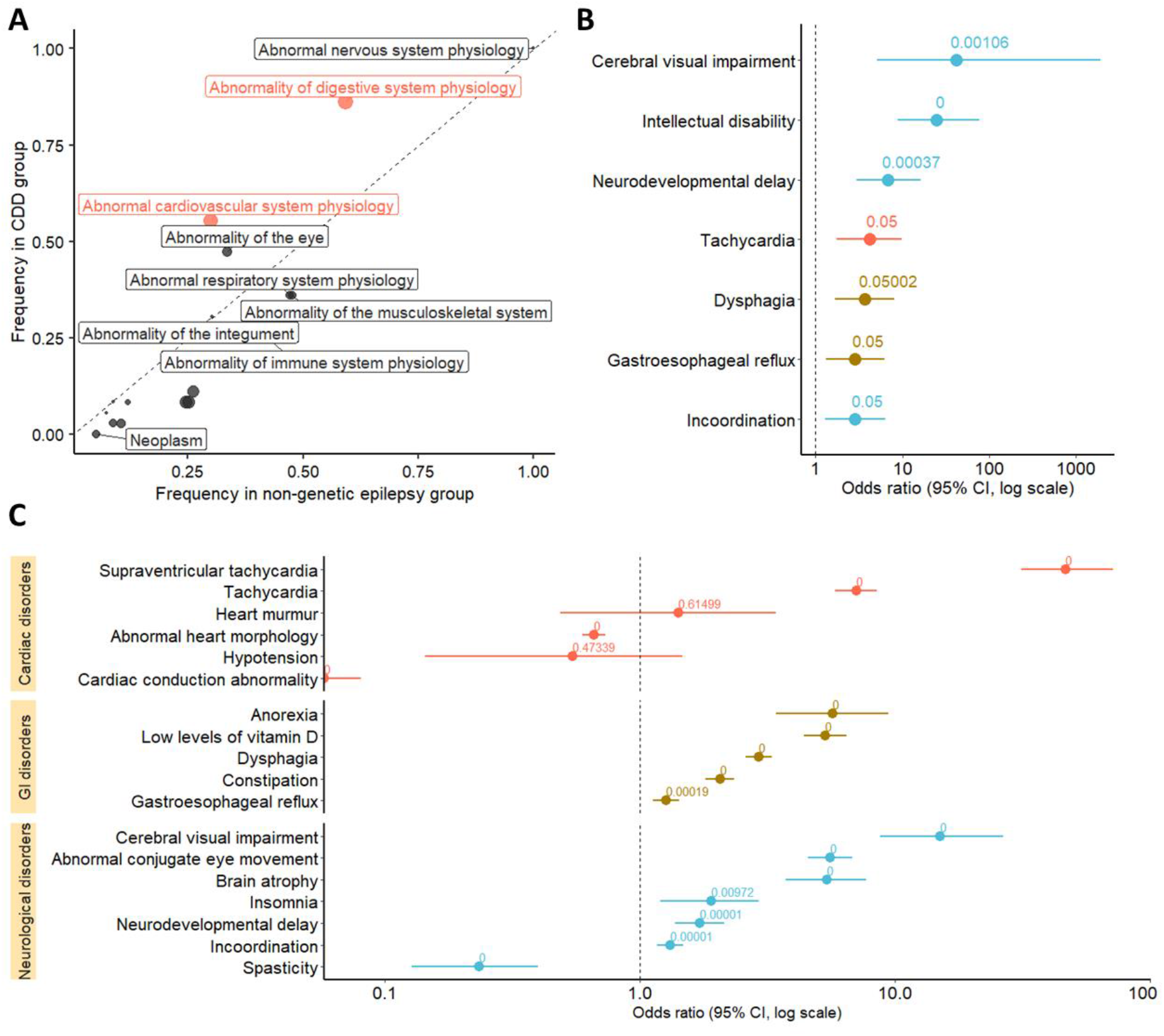
Human Phenotype Ontology (HPO) term enrichment in CDKL5 deficiency disorder in comparison to individuals with epilepsy. **A**. Frequency of body systems abnormalities coded as Human Phenotype Ontology (HPO) terms in CDD and matched control group. Statistically significant difference is colored red. Abnormalities of eye, cardiovascular system, and digestive system had a higher frequency in CDD patients; however, only abnormality of digestive system physiology (HP:0025032) and abnormal cardiovascular system physiology (HP:0011025) reached statistical significance (p=0.03 for both). The terms plotted above the dashed line are more frequent in individuals with CDD. Terms in which the frequency difference reached statistical significance after adjustment for multiple comparisons are colored red. **B**. Forest plot of individual-based specific HPO term enrichment that reached statistical significance after adjustment for multiple comparisons. Annotated are adjusted P values associated with a particular term. Cerebral visual impairment (HP:0100704), intellectual disability (HP:0001249), neurodevelopmental delay (HP:0012758), incoordination (HP:0002311), dysphagia (HP:0002015), and gastroesophageal reflux (HP:0002020) are well-described CDD comorbidities. Tachycardia (HP:0001649) was significantly enriched in CDD cohort along with established comorbidities. **C**. Forest plot of encounter-based concept enrichment for HPO terms characterizing CDD phenotype sorted by systems and arranged by size effect. Annotated are adjusted P values associated with a particular term. Medical encounter notes of individuals with CDD were significantly enriched for tachycardia (HP:0001649), as well as supraventricular tachycardia (HP:0004755). Abbreviations: CDD, CDKL5 deficiency disorder; HPO Human Phenotype Ontology.

To obtain more clinical information for the cardiovascular findings from the EHR screening, we performed manual chart review (Supplementary Table 2). We found that 18 out of 38 individuals with CDD had tachycardia in at least one encounter unrelated to febrile illness. People with CDD and tachycardia were 5.63 times more likely to also have other autonomic symptoms, such as drooling or urinary retention, compared to individuals with CDD who did not have tachycardia (95%CI 1.08-40.3, p=0.038). Of 38 individuals, 16 had ECGs available and only for 5 of them (31%) tachycardia was captured in the ECG performed at a respective encounter. Two individuals had history of fetal/neonatal SVT, which has never been reported in CDD; their cases are described in Table 1. One person had long QT interval, which was not confirmed by subsequent ECGs. One individual had an atrial septal defect, which was not more prevalent than in control group (OR 1.25, 95%CI 0.02-13.16, p_adj_=1).

**Table 1.** Clinical description of fetal/neonatal supraventricular tachycardia in individuals with CDD.

| ID | Variant | GA, weeks | Gender | Description |
| --- | --- | --- | --- | --- |
| 10 | NM_003159.2:c.351T>A (p.Tyr117*) | 39 | Male | Developed SVT at the heart rate of 210 beats/minute at 36 hours of age. ICU monitoring showed SVT with an abundance of premature ventricular contractions. The patient was administered adenosine without success and subsequently cardioverted. He was started on propranolol and discharged with normal cardiac rate and rhythm. Treatment with propranolol was stopped at 1.5 months of age. The patient was further noted to have tachycardia. |
| 12 | NM_003159.2: c.2323_2326del (p.Glu775Serfs*8) | 39 | Male | Born via C-section due to abnormal heart rhythm and diagnosed with SVT at 290 beats/minute after being born. SVT was medically aborted and the patient was discharged from the neonatal ICU on atenolol and flecainide. Both medications were first discontinued at 6 months but recommenced several times over the patient's lifetime because of SVT recurrence. |

Abbreviations: GA, gestational age; SVT, supraventricular tachycardia.

## Discussion

Using EHR data analysis augmented by automated HPO annotations in the setting of *CDKL5* Center of Excellence, for the first time, we replicated previous findings in a computational analysis of real-world data. With this approach, we also showed that prevalence of tachycardia is higher in CDD patients than in matched controls with pediatric-onset epilepsy. Although tachycardia was confirmed by ECG in minority of cases, we noted that it is more prevalent in individuals with other concurrent autonomic symptoms. The computational term-frequency analysis in encounter notes against matched controls allowed us to identify two cases of fetal/neonatal SVT not previously described in CDD. Cardiovascular symptoms are not universal in individuals with CDD in our cohort, which might be attributed to the known phenotypic variability of CDD and could be addressed in the future genotype-phenotype studies.

The computational approach we used could validate existing CDD findings and discover potential novel associations that can be further explored in further studies. Computational phenotyping offers numerous advantages for research design and conductance, including faster pipeline, reduced costs, and the ability to examine a wide range of phenotypes. This approach is extendable to other rare diseases, as demonstrated by previous research in the field.^15,16^ In the future, it can be employed for large-scale studies on rare diseases with integration of data from various healthcare systems similar to that conducted currently in common disease research.^17,18^

Cardiovascular symptoms, including QT interval are a recognized part of phenotype in Rett syndrome^5^, a closely related condition, confirmed by animal model studies.^19^ Likewise, a recent study demonstrated lQT and tachycardia in a mouse model of CDD.^6^ In 47% of our cohort in cross-sectional analysis clinical tachycardia was noted. Although it was not seen on routine ECG in most cases, none of our patients had a long-term cardiac monitoring, which would be more feasible to confirm this finding. We hypothesize that tachycardia in CDD could be a manifestation of dysautonomia. CDD animal model studies have shown altered cholinergic drive^20^ and decreased vagal output to the heart.^6^ Clinically, individuals with CDD often present with other dysautonomic features, such as gastrointestinal motility issues and respiratory abnormalities^11^, which we also observed in our study.

The occurrence of fetal/neonatal SVT in CDKL5 deficiency disorder has not been previously described. Fetal SVT warrants immediate delivery or intrauterine antiarrhythmic treatment as it may result in fetal hydrops and intrauterine demise. Neonatal SVT may result in morbidity if left untreated and bears an increased risk of persistent tachycardia. Several types of SVT according to the disease mechanism exist; however, due to the retrospective nature of the study, we could not confirm the exact type of SVT, which limits the pathophysiological interpretation of the finding.

The findings from our study contribute to understanding of CDD comorbidity spectrum and give rationale for further clinical characterization of cardiovascular features in individuals with CDD. The current consensus recommendations for management of individuals with CDD suggest obtaining a baseline ECG and cardiovascular screening but not a longitudinal follow-up, noting that there is lack of data on cardiac arrhythmias in individuals with CDD.^8^ Pending further validation, our study may inform future guidelines regarding cardiac surveillance in CDD.

Our study has several limitations. Electronic health records are real-world data that are not collected for research purposes. This data source is subject to the data quality of clinical notes and billing codes, and susceptible to multiple sources of bias. The mapping procedure across the UMLS and HPO is established^14^, but this mapping is incomplete and may lead to underrepresentation of certain phenotypic features.

In conclusion, we applied a novel approach screen a rare genetic disorder on healthcare network scale, validated previous research findings in real-world data computationally, and identifies tachycardia and fetal/neonatal SVT as potentially novel phenotypes associated with CDD, which is in line with data from animal model^6^ and warrants accounting for cardiovascular phenotype in the future large multicenter phenotyping studies. The presented approach is fast, resource-efficient, and scalable to healthcare systems and large-scale retrospective studies that inform follow-up targeted clinical studies.

## Data Availability

The data supporting the findings of this study are available within the article and its supplementary material.

## Data Availability

The authors confirm that the data supporting the findings of this study are available within the article and its supplementary material.

## Author Contributions

Conceptualization: D.L., E.P.K., A.I.; Supervision: D.L., E.P.K.; Data Curation: E.P.K., A.I., P.A., X.Z.; Methodology: A.M., M.J., S.T., G.K., A.I., C.B., D.L.; Visualization: A.I., C.B., S.T.; Formal Analysis: A.I., Writing-original draft: A.I.; Writing-review and editing: D.L., E.P.K., A.I., A.M., M.J., S.T., G.K., A.I., C.B., P.A., X.Z.

## Ethics Declaration

This study was approved by IRB, approval ID 22-147. Informed consent was waived considering the retrospective nature of the study.

