## Supplemental Material for "Natural language processing and expert follow-up establishes tachycardia association with *CDKL5* deficiency disorder"

### Supplementary files

Alina Ivaniuk[1,2], Christian M Boßelmann[1,2], Xiaoming Zhang[1,2], Mark St John[1,2], Sara C Taylor[3], Gokul Krishnaswamy[3], Alex Milinovich[4], Peter F Aziz[5], Elia Pestana-Knight[2], Dennis Lal[1,3,6,7]

[1]Genomic Medicine Institute, Lerner Research Institute, Cleveland Clinic, Cleveland, OH, USA.

[2]Epilepsy Center, Neurological Institute, Cleveland Clinic, Cleveland, OH, USA

[3]Neurological Institute, Cleveland Clinic, Cleveland, OH, USA

[4]Department of Quantitative Health Sciences, Cleveland Clinic, Cleveland, OH, USA

[5]Department of Pediatric Cardiology, Cleveland Clinic, Cleveland, OH, USA

[6]Stanley Center for Psychiatric Research, Broad Institute of Harvard and M.I.T., Cambridge, MA, USA.

[7]Cologne Center for Genomics (CCG), University of Cologne, Cologne, Germany

### Supplementary Figure 1. Cohort construction

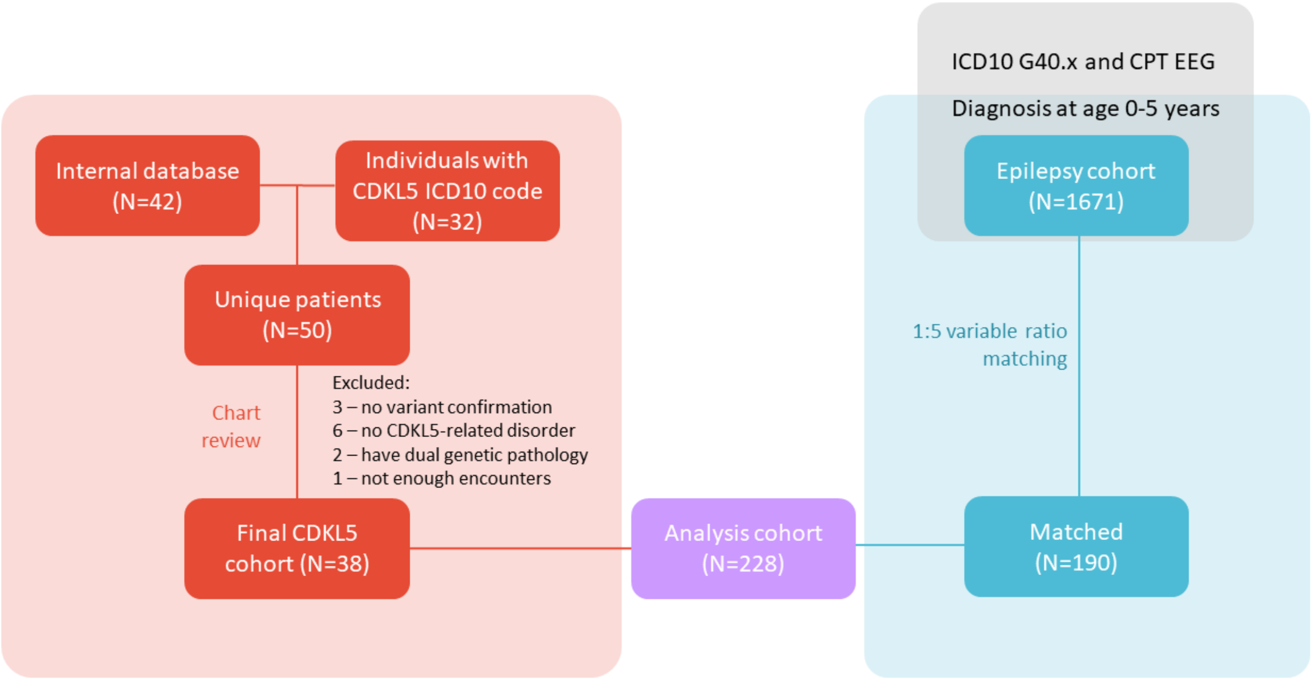

**Figure S1.** We included 42 people with CDKL5 deficiency disorder (CDD) listed in the internal database of the CDKL5 Center of Excellence at Cleveland Clinic Main Campus in Cleveland, Ohio. To ensure complete inclusion of individuals with CDD from other clinics of the Cleveland Clinic healthcare network, we additionally screened network-wide health records for the ICD-10 code G40.834 (Cyclin-Dependent Kinase-Like 5 Deficiency Disorder). The computational search identified eight additional patients. For the combined cohort of 50 people with CDD, we performed electronic chart review to verify the genetic diagnosis. Three individuals without genetic variant confirmation, six individuals who were coded with an ICD-10 code for CDD but turned out not to have the disorder, two having a concurrent pathogenic variant in another gene (*SCN1A*), and one patient for having only 1 encounter in the system were removed from the analysis. To construct the matched control cohort, we first identified 1671 individuals with epilepsy by querying the health record database for patient identifiers satisfying the following criteria: a) Having any ICD-10 G40 ("Epilepsy and recurrent seizures") or ICD-9 345 code; b) Having a Current Procedural Terminology (CPT) code for any type of electroencephalography (EEG); c) Age 0-5 years at time of diagnosis (defined as an age of the first billing code for epilepsy). After nearest neighbor propensity score matching (see methods), we derived the matched control cohort of 190 individuals.

### Supplementary Figure 2. UMLS mapping process

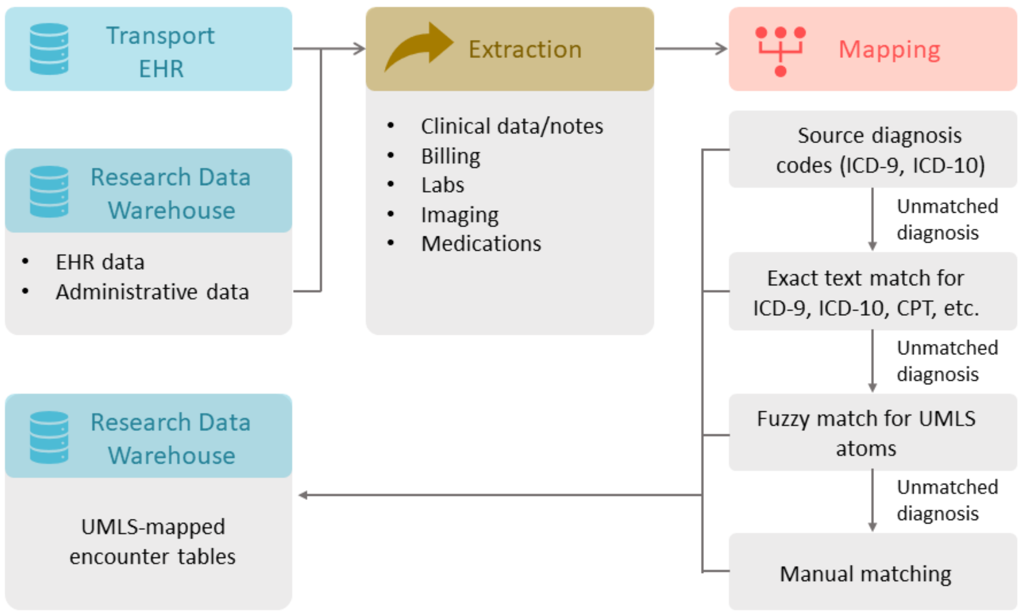

**Figure S2.** Research Data Warehouse (RDW) is an in-house relational database that hosts encounter tables mapped to Unified Medical Language System (UMLS, release 2022AA) concepts from different data sources. The data sources include electronic health record (HER; in this case, Epic) entries and administrative data sources hosted within RDW itself, as well as transport EHR (in this case, Golden Hour/emsCharts). Extracted data, which includes clinical notes, billing codes, laboratory and imaging results, and medication data is mapped to UMLS in several steps. At the first step, the source diagnosis codes, either ICD-9 or ICD-10, are directly mapped to UMLS. If the raw data entity does not include an associated diagnosis code, the next step involves mapping through exact unstructured text matching to same-source codes. Unstructured text, e.g. encounter notes, were analyzed with cTAKES[1], a natural language processing software, to extract medical phrases for mapping to UMLS and HPO. Remaining unmatched data entities undergo fuzzy matching for UMLS atoms, i.e., the smallest units of naming within UMLS system. All remaining unmatched data entities are manually mapped. Detailed description of database development is given by Reimer and Milinovich, 2020.[2]

### Supplementary Figure 3. Human Phenotype Ontology term propagation

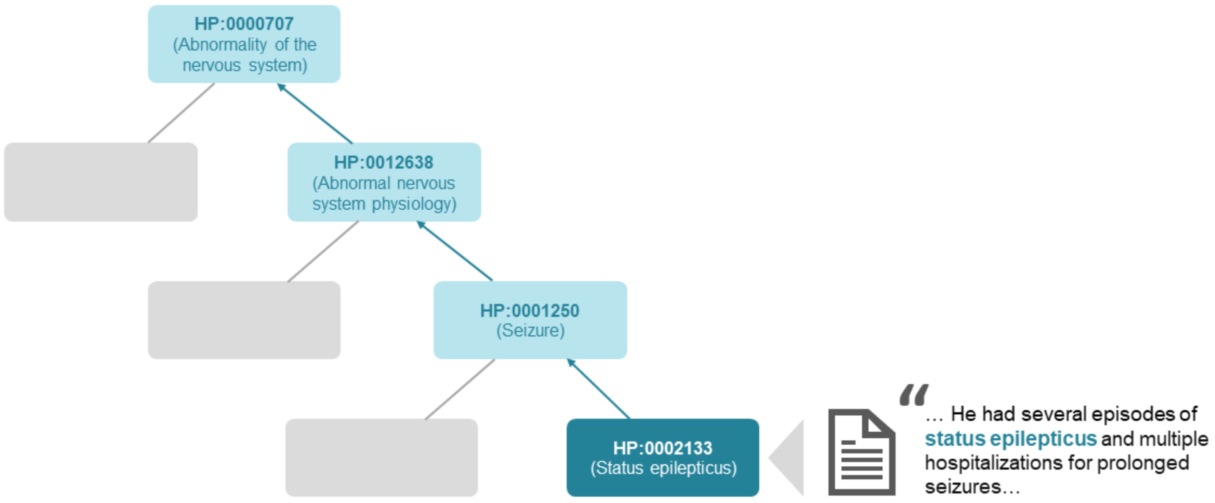

**Figure S3.** Human Phenotype Ontology (HPO) is a comprehensive hierarchical biomedical term library in which each term represents a more specific instance of a corresponding parent term. Propagation refers to identification of all parent terms of a particular term. Propagation facilitates phenotypic data harmonization and allows for phenotype exploration on different levels of specificity.

### Supplementary Table 1. CDD and matched control cohort comparison

| Variable | | CDD | Control | P |
|---|---|---|---|---|
| Gender | Female, n(%) | 28 (73.7) | 132 (69.5) | 0.746 |
| | Male, n(%) | 10 (26.3) | 58 (30.5) | |
| Ethnicity | Hispanic or Latino, n(%) | 3 (7.9) | 14 (7.4) | 1.000 |
| | Not Hispanic or Latino, n(%) | 35 (92.1) | 176 (92.6) | |
| Number of encounters | Median (IQR) | 38.0 (14.0 to 114.0) | 22.0 (9.0 to 85.5) | 0.357 |
| Minimum encounter age | Median (IQR) | 0.2 (0.0 to 1.1) | 0.2 (0.0 to 1.3) | 0.674 |
| Median encounter age | Median (IQR) | 3.4 (1.6 to 7.5) | 4.3 (2.1 to 7.0) | 0.630 |
| Maximum encounter age | Median (IQR) | 11.6 (6.5 to 16.3) | 9.1 (4.9 to 19.1) | 0.762 |
| Length of follow-up | Median (IQR) | 11.3 (5.0 to 16.0) | 8.2 (2.6 to 19.1) | 0.882 |

### Supplementary Table 2. Clinical characteristics and cardiovascular phenotype of CDD cohort

| ID | Sex | Variant | ACMG/AMP classification | Cardiac phenotype description | Autonomic features | Stimulant medications | Ambulatory status |
|---|---|---|---|---|---|---|---|
| 1 | Female | NM_003159.2:c.668T>G (p.Ile223Ser) | LP | Tachycardic, short PR interval | Drooling, irregular breathing | No | No |
| 2 | Female | NM_003159.2:c.529T>C (p.Tyr177His) | LP | Tachycardic | Drooling, urine retention, cold extremities | No | With support |
| 3 | Female | NM_001323289.2:c.2322del (p.Glu775Argfs*9) | P | Tachycardic | Drooling, cold extremities | No | No |
| 4 | Male | NC_000023.11(NM_003159.3):c.99+5G>A (splice) | P | Hypertension and cardiomyopathy after treatment with ACTH for epileptic spasms. Deceased at teenage following cardiac arrest. | Drooling | No | No |
| 5 | Female | NM_003159.2:c.1498_1501dup (p.Leu501Glnfs*4) | LP | Tachycardic | Drooling | No | With support |
| 6 | Female | NM_001323289.2:c.2374dup (p.Thr792Asnfs*9) | LP | Tachycardic | Drooling, cold | No | No |

| | | | | | | | |
|---|---|---|---|---|---|---|---|
| | | | | | extremities, urinary retention | | |
| 7 | Female | NM_001323289.2:c.533G>T (p.Arg178Leu) | P | Tachycardic | None | No | No |
| 8 | Female | NC_000023.11:g.18582354_18626826del (deletion including exons 4-12) | P | Tachycardic | Drooling | No | Yes |
| 9 | Female | NM_001323289.2:c.533G>A (p.Arg178Gln) | P | ASD closed at teenage; Tachycardic | Drooling, cold feet | No | No |
| 10 | Male | NM_003159.2:c.351T>A (p.Tyr117*) | P | Neonatal SVT. Treated with propranolol until the 1.5 months of age. No SVT recurrence after discontinuation of propranolol. Hypertention due to treatment with ACTH. Tachycardic. | Drooling, cold feet | No | Yes |
| 11 | Female | NM_003159.2:c.623del (p.Gln208Argfs*20) | P | None | Drooling | Yes | Yes |
| 12 | Male | NM_003159.2:c.2323_2326del (p.Glu775Serfs*8) | P | C-section due to abnormal fetal rhythm (SVT) treated with beta-blockers and flecainide; PFO | Drooling, cold extremities | No | No |

| | | | | | | | |
|---|---|---|---|---|---|---|---|
| 13 | Female | NM_003159.2:c.784T>C (p.Tyr262His) | LP | Tachycardic | Cold feet | No | No |
| 14 | Female | NM_001323289.2: c.364G>A (p.Ala122Thr) | P | Tachycardic, PFO | Drooling | No | No |
| 15 | Female | NM_001323289.2:c.1675C>T (p.Arg559*) | P | Tachycardic | Drooling, cold extremities | No | No |
| 16 | Male | NM_001323289.2:c.1976_1977del (p.Val659Glyfs*23) | P | Sinus bradycardia, followed by tachycardia | None | No | Yes |
| 17 | Male | NM_001323289.2:c.1165C>T (p.Gln389*) | P | Tachycardic and lQT syndrome. Died due to cardiorespiratory compromise. | Drooling, cold extremities, irregular breathing | No | No |
| 18 | Female | NC_000023.11(NM_003159.3):c.65-1G>A (splice) | P | Tachycardic | Cold feet | No | Yes |
| 19 | Female | NM_001323289.2:c.1354C>T (p.Gln452*) | P | Tachycardic | Drooling, cold extremities | No | No |
| 20 | Female | NM_001323289.2:c.629del (p.Leu210Tyrfs*18) | P | Bradycardia, hypertension, LVH due to ACTH treatment for epileptic spasms; Tachycardic | None | No | With support |
| 21 | Female | NM_001323289.2:c.175C>T (p.Arg59*) | P | Tachycardic, PFO, right ventricular hypertrophy | None | Yes | No |

| | | | | | | | |
|---|---|---|---|---|---|---|---|
| 22 | Female | NM_001323289.2:c.595T>C (p.Cys199Arg) | LP | Left ventricular hypertrophy | None | No | No |
| 23 | Male | NM_001323289.2:c.400C>T (p.Arg134*) | P | None | Irregular breathing, cold feet, drooling | No | Yes |
| 24 | Female | NC_000023.11(NM_003159.3):c.2046+1G>A (splice) | LP | None | Drooling, cold feet | No | No |
| 25 | Male | NM_001323289.2:c.215T>C (p.Ile72Thr) | P | None | None | No | No |
| 26 | Male | NM_001323289.2:c.608A>G (p.Glu203Gly) | LP | None | Drooling | No | No |
| 27 | Female | NM_001323289.2:c.412C>G (p.Pro138Ala) | P | None | None | No | No |
| 28 | Female | NM_001323289.2:c.470C>T (p.Ala157Val) | LP | None | None | No | No |
| 29 | Male | NM_001323289.2:c.587C>G (p.Ser196Trp) | LP | None | Drooling | No | No |
| 30 | Female | NM_001323289.2:c.934A>T (p.Leu312*) | P | None | Irregular breathing | No | Not applicable (too young) |
| 31 | Female | NM_001323289.2:c.46del (p.Val18Leufs*2) | P | None | None | No | Not applicable (too young) |
| 32 | Female | NM_001323289.2:c.725del (p.Pro242Leufs*25) | P | None | None | No | No |
| 33 | Female | NC_000023.11:g.18209153_19223560del | P | None | None | No | No |
| 34 | Female | NC_000023.11(NM_001323289.2):c.65-2A>T | P | None | Cold feet | No | No |

| | | | | | | | |
|---|---|---|---|---|---|---|---|
| 35 | Female | NM_001323289.2:c.1244_1245del (p.Thr415Argfs*3) | LP | None | Drooling | No | No |
| 36 | Female | NM_001323289.2:c.1988del (p.Ser563Thrfs*121) | LP | None | Drooling | No | No |
| 37 | Male | NM_001323289.2:c.103A>C (p.Thr35Pro) | LP | None | None | No | No |
| 38 | Female | NC_000023.11:g.18564468_18564474delinsGCAGA (splice) | P | None | Drooling | No | Yes |

Abbreviations: ACTH, adrenocortocotropic hormone; ASD, atrial septal defect; ECG, electrocardiogram; LP, likely pathogenic; LVH, left ventricular hypertrophy; lQT, long QT syndrome;  P, pathogenic; PFO, patent foramen ovale; SVT, supraventricular tachycardia.
